# Prevalence, pattern, and factors associated with ocular morbidity among stone quarry workers in districts of Kampala and Mukono, Uganda

**DOI:** 10.64898/2026.01.26.26344885

**Authors:** Ahebwa Amelia, Otiti Juliet, Atukunda Immaculate, Ntende Jacob, Ssali Grace, Abdalla Ahmed, Moses Kasadhakawo, Lusobya Rebecca Claire, Ampaire A. Musika, Kamya Frank, Sempa Lwanga, Mugabi Barnabas, Agaba Marianna

## Abstract

**Objective:** To determine the prevalence, pattern, and associated factors of ocular morbidity among stone quarry workers in Kampala and Mukono, Uganda.

**Methods and Analysis:** In a field-based cross-sectional study, we recruited 290 stone quarry workers using consecutive and simple random sampling. A pretested questionnaire was used to collect relevant information on patient history and ocular examination findings. Ocular morbidity was defined as any abnormal finding on eye examination. Associated factors were analyzed using logistic regression with odds ratios and 95% confidence intervals.

**Results:** The mean age of the participants was 35 years (SD +/-12). The prevalence of ocular morbidity was found to be as high as 80%. The common ocular disorders found included pinguecula, pterygium, presbyopia, conjunctival hyperpigmentation, and dry eye. There was a statistically significant relationship between ocular morbidity and the following factors: age over 40 years, type of work engaged at the quarry, duration of 5 years and greater working at the quarry, inconsistent usage of protective eyewear and no observation of any workmate suffering vision-threatening eye injuries.

**Conclusion:** The prevalence of ocular morbidity was very high at 80%, with the most common morbidities being ocular surface disorders. The associated factors included age, type of work performed at the quarry, duration of working at the quarry, use of protective eyewear, and lack of observation of any workmate suffering vision-threatening eye injuries. Main limitations included potential recall bias in regards to self-reported occupational accidents, and lack of pre-employment ocular health records for baseline comparison.

**Key messages^1^:** *What is already known on this topic:* Work-related ocular injuries are a major global public health concern and have been prioritized by the WHO and ILO due to their impact on health, productivity and achievement of the Sustainable Development Goals. However, occupational ocular morbidity among stone quarry workers in Uganda is poorly documented, creating a critical evidence gap that this study sought to address to inform occupational safety and eye health policy.

*What this study adds:* This study provides the first evidence from Uganda demonstrating a very high burden of ocular morbidity (80%) among stone quarry workers, with conjunctival disorders and presbyopia being the most common conditions. It also identifies key occupational and behavioral factors associated with ocular morbidity, including older age, high-risk quarry tasks, longer duration of work, inconsistent use of protective eyewear and lack of exposure to severe eye injury events.

*How this study might affect research, practice or policy:* The findings highlight the urgent need for improved access to and proper use of eye protective equipment, and strengthened enforcement of occupational safety regulations in quarry settings. These results can inform occupational health policy, guide targeted preventive interventions, and serve as baseline data for future research in similar low-resource settings.

## INTRODUCTION

According to a joint report by the International Labor Organization (ILO), International Agency for the Prevention of Blindness (IAPB), and World Health Organization (WHO), ocular morbidity remains a major public health problem and an important cause of preventable visual impairment globally, with an estimated 13 million people living with vision impairment linked to their work and an estimated 3.5 million eye disorders occurring in the workplace annually [1]. Compared with those in developed countries, occupational eye disorders account for 21% of all workplace injuries of workers in mining, construction, and manufacturing, who are particularly at risk, with a higher prevalence in low- and middle-income countries (LMICs); this difference is attributable to disparities in the availability of eye safety devices, safety and health training, and regular workplace supervision [2].

Stone quarrying is a particularly hazardous occupation with the potential for ocular morbidity arising from work-related activities such as cutting, hammering, drilling, blasting, and the use of pneumatic chisels for the excavation of stones used in building and making roads. The environment of quarry activities is also dry, dusty and heavily polluted [3]. The use of rudimentary tools and noncompliance with the use of personal protective equipment increases the risk of projectile chips of stone, metal, and wood entering the eyes, thus resulting in corneal lacerations and penetrating ocular injuries. Other common injuries include destabilization of the pre-corneal tear film, radiation-induced conjunctival degeneration and lens changes, superficial ocular surface abrasions and foreign bodies [4].

Ocular morbidity and vision impairment in the quarry industry often occur as a consequence of work-related hazards that precipitate pathological processes or aggravate preexisting eye pathology. Work-related ocular morbidity among stone quarry workers subsequently leads to blindness, visual impairment, work absenteeism, suboptimal work output, and injury-related compensation claims [3]. Furthermore, work-related eye injuries alone cost more than $300 million per year due to loss of production time, medical treatment, and worker compensation [5].

The Occupational Safety and Health (OSH) Act 2006 section 19 streamlines the need for employers to provide PPE for workers and ensure usage while at the workplace; however, this is not the case, as evident from several studies, including a recent study in Uganda that revealed that existing OSH laws were largely outdated compared to the current needs of workplaces. The Ugandan-based study also pointed to challenges affecting implementation arising from gaps in the legal framework, low public awareness about OSH, poor planning, limited human capacity, transparency, and accountability [6].

Wearing adequate protective eyewear is the most important thing for the protection of vision at work and can prevent more than 90% of serious work-related ocular pathologies [7]. This suggests the need for adequate ocular protection during quarrying activities and appropriate occupational health measures to ensure the optimum oculo-visual health and well-being required for productivity [8].

Therefore, this study explored ocular health status among stone quarry workers by assessing the prevalence, pattern, and factors associated with ocular morbidity among the selected population in the districts of Kampala and Mukono, Uganda. The findings from the study will serve as a baseline for further research and subsequent policy development aimed at improving the occupational eye health of workers in the stone quarry industry in Uganda and other low-income countries.

## MATERIALS AND METHODS

### Study Design and Setting

This was a field-based cross-sectional study conducted from February to March 2024 in all the operational stone quarries located in the Bukasa and Nakasajja areas of the Kampala and Mukono districts, respectively. The two districts have a significant number of functional quarries serving metropolitan areas that are characterized by high demand and supply of stone for construction and architectural purposes. Bukasa is located in the Makindye division and is approximately 8 km from Kampala’s central business district. Bukasa is a commercial place where all kinds of businesses and activities take place, including the lucrative stone mining business, but it also doubles as a residential area. Nakasajja is located in the Mukono district within the central region of Uganda. It is bordered by Kabubu in the North, Kitukutwe in the South, Nakwero in the West, and Kasambya in the East.

### Study Population and Sampling

This study was conducted among stone quarry workers exposed to occupational hazards during the excavation and processing of stones in quarry sites within Bukasa, Kampala city, and Nakasajja, Mukono district, during the study period (February 2024-March 2024). Participation in the study was voluntary, and informed consent was obtained from the study participants. Unwell participants were excluded. Using the Kish–Leslie equation for cross-sectional studies, a sample size of 290 participants was obtained for the study. Consecutive sampling coupled with simple random sampling was performed to select the study participants who fulfilled the eligibility criteria.

### Data collection and procedures

This involved the use of a face-to-face interviewer-administered questionnaire for study participants following screening. The administration of the pretested structured questionnaire, screening, and data collection were performed by the Principal Investigator and trained Research Assistants (Ophthalmic clinical officers and nurses). The data included socio-demographic data (age, sex, educational background, marital status), medical information (history of trauma, the agent causing injury, use of spectacle correction, history of eye surgery, history of use of ocular medication), occupational and social history (nature of work, pre-employment eye examination) and clinical eye examination findings.

The eye examinations included distance visual acuity using a 6 m Snellen’s chart or illiterate E chart; those with visual acuity (V/A) less than 6/6 were reassessed with a pinhole and underwent refraction. Near visual acuity was assessed using a Jaeger chart, and refraction was performed on all participants with impaired near vision. Visual fields were assessed by the confrontational method in comparison with the examiner (the examiner had normal visual fields confirmed by perimetry). Extra-ocular muscle movements were assessed, and diplopia was sought in all directions of gaze. Extra-ocular muscle activity was also assessed using the cover-over uncover test to assess phorias. An Amsler grid was used in all subjects to assess macular function. The lids, conjunctiva, cornea, anterior chamber, pupil, and iris were examined using a portable slit lamp. Tonometry using an I-care tonometer was performed for all participants. In study participants with visual acuity less than 6/6, dilatation of the pupil was performed using tropicamide/phenylephrine eye drops, and then direct ophthalmoscopy was performed.

### Statistical analysis

The data collected from the studies were entered into EpiData version 4.1 and then exported to STATA version 14.0 for data cleaning and analysis. Continuous variables were summarized using means and standard deviations for normally distributed data and medians and percentiles (75th and 25th) for non-normally distributed data. Categorical variables are summarized as proportions and percentages. The results are presented in tables and graphs.

The prevalence of ocular morbidities in stone quarry workers was calculated as the proportion of those with any ocular abnormalities (numerator) among the total number of stone quarry workers in the study (denominator) and expressed as a percentage. Additionally, 95% confidence intervals were estimated. The various ocular abnormalities were summarized using proportions, percentages, and graphs.

To explore factors associated with ocular abnormalities, bivariate and multivariate analyses were performed using logistic regression methods. Factors with a p-value <0.2 in the bivariate analysis were considered for inclusion in the multivariate analysis. Confounding and interaction effects were assessed via multivariate analysis. Factors with p values less than 5% in the multivariable analysis were considered statistically significant.

## RESULTS

### Baseline characteristics of the study population

We recruited a total of 290 participants with an age range of 16 to 72 years and a mean age of 35 years (SD +/-12). There were 175 (60.3%) males and 115 (39.7%) females, for a male-to-female ratio of 2:1. The majority of the stone quarry workers (62.1%) were Breakers and Crushers. There were 250 (86.2%) participants who reported not using PPE (goggles or helmets), while 169 (58.3%) had a history of ocular injury. More than half of the quarry workers (159, 54.8) had worked for less than 5 years at the quarry (study site), and more than three-quarters (221, 76.2) had worked more than 10 hours per day (Table 1).

**Table 1:** Socio-demographic and occupational characteristics of the interviewed stone quarry workers.

| Variable | Categories | n (%) |
| --- | --- | --- |
| Age (in completed years) | Less than 18 years | 27(9.3) |
|  | 18 -28 years | 88(30.3) |
|  | 29 - 39 years | 80(27.6) |
|  | 40 years and above | 95(32.8) |
| Sex | Male | 175(60.3) |
|  | Female | 115 (39.7) |
| **Education** |  |  |
|  | None | 36(12.4) |
|  | Primary | 127(43.8) |
|  | Secondary | 127(43.8) |
| **Marital status** |  |  |
|  | Single | 113(39.0) |
|  | Married | 163(56.2) |
|  | Widowed/Divorced | 14(4.8) |
| **Religious denomination** |  |  |
|  | Christian | 232(80.0) |
|  | Muslim | 58(20.0) |
| **Type of work done at the quarry** |  |  |
|  | Blasters/drillers | 16 (5.5) |
|  | Laborers | 59 (20.3) |
|  | Breakers | 35 (12.1) |
|  | Crushers | 180 (62.1) |
| **Duration of work at the quarry** |  |  |
|  | Less than five years | 159 (54.8) |
|  | Five years and above | 131 (45.2) |
| **Use of any spectacle correction** |  |  |
|  | Yes | 8(2.8) |
|  | No | 282 (97.2) |
| **Use of protective eyewear** |  |  |
|  | Yes | 40 (13.8) |
|  | No | 250 (86.2) |
| **Type of eye personal protective equipment used** |  |  |
|  | Goggles | 10 (25.0) |
|  | Helmet | 30 (75.0) |
| **Frequency of using eye personal protective equipment** |  |  |
|  | Always | 10 (25.0) |
|  | Sometimes | 19 (47.5) |
|  | Rarely | 11 (27.5) |
| **Number of work days per week** |  |  |
|  | Three days | 53 (18.3) |
|  | Four days | 80 (27.6) |
|  | Five days | 114 (39.3) |
|  | More than five days | 43 (14.8) |
| **Work duration in quarry per day** |  |  |
|  | Less than 10 hours | 69 (23.8) |
|  | More than 10 hours | 221 (76.2) |
| **Tools usually used daily in quarry work** |  |  |
|  | Regular hammer (Rounded bottom tip) | 7 (2.4) |
|  | Regular hammer (Flat bottom) | 89 (30.7) |
|  | Sledge hammer | 46 (15.9) |
|  | Pick axe | 67 (23.1) |
|  | All the above | 81 (27.9) |
| **Stones majorly produced** | Quarter inch only | 42(14.5) |
|  | Half inch only | 72(24.8) |
|  | Hardcore only | 32(11.0) |
|  | Quarter-inch and half-inch | 144(49.7) |

### Prevalence of ocular morbidity

In this study, the prevalence of ocular morbidity among stone quarry workers in the districts of Kampala and Mukono, Uganda, was 80% (95% CI: 75.26-85.63), which is four-fifths of the population of workers interviewed (Table 2).

**Table 2:** Prevalence of ocular morbidity among stone quarry workers.

|  | *Frequency* | *Prevalence* | *[95% Conf. Interval]* |
| --- | --- | --- | --- |
| *No ocular disorder* | 57 | 20% | 14.57-24.94 |
| *Ocular disorder* | 233 | 80% | 75.26-85.63 |

### Pattern of ocular morbidity

The most common ocular morbidities were conjunctival disorders, particularly pinguecula (111, 38.2%) and pterygium (99, 34.1%), followed by presbyopia (82, 28.2%), conjunctival hyperpigmentation (60, 20.6%), dry eye (28, 9.6%) and cornea disorders. Several vision-threatening abnormalities were also present, including refractive errors, eyelid abnormalities, corneal disorders, cataracts and retinal disorders (Fig. 1).

**Figure 1:**
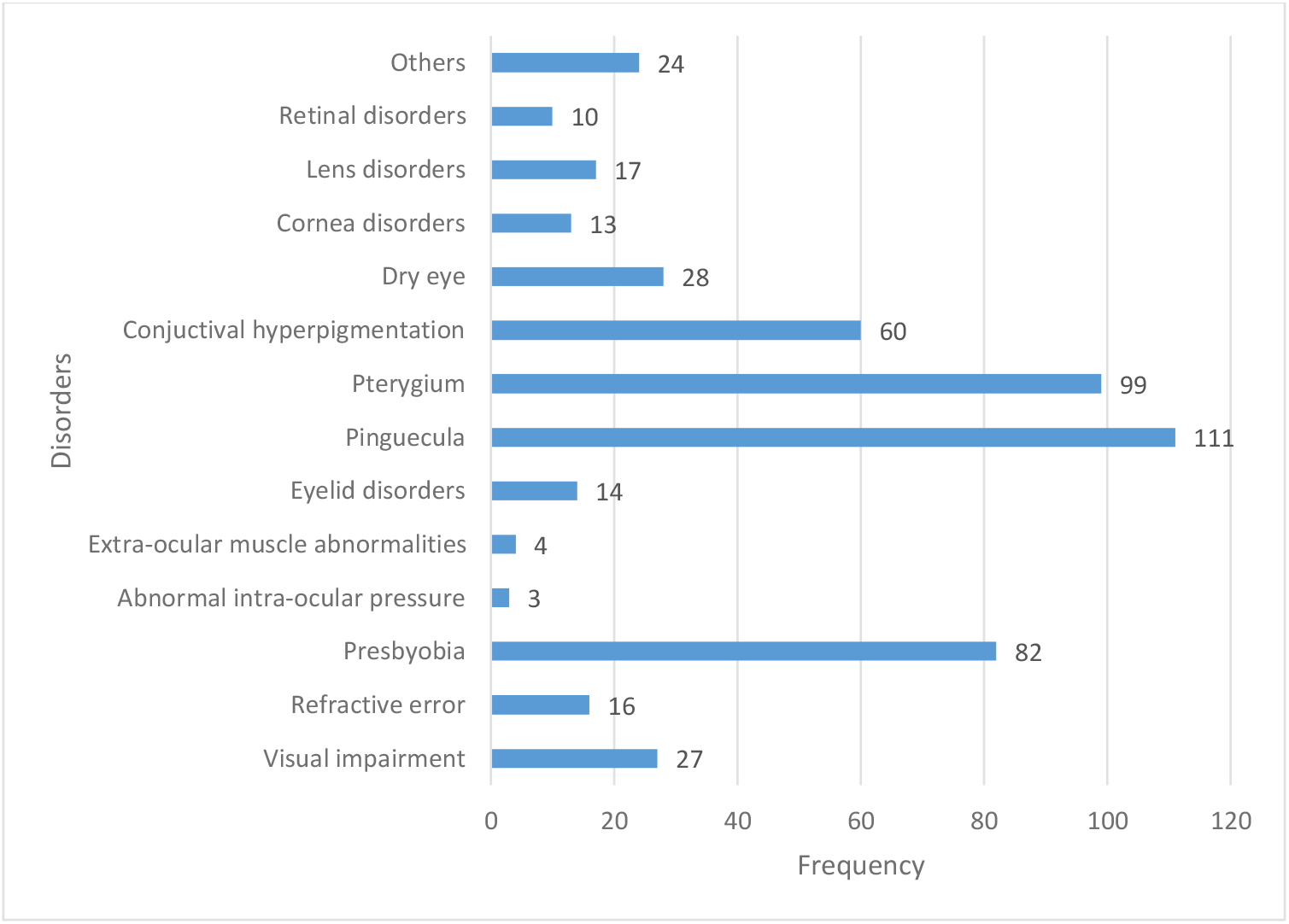
Number of participants with different ocular disorders.

### Factors associated with ocular morbidity among stone quarry workers

Five factors showed statistically significant associations with ocular morbidity at the bivariate level (p<0.2). This included age, the type of work engaged at the quarry, the duration of working at the quarry (study site), the use of protective eyewear, and non-observation of any workmate suffering vision-threatening eye injuries. When the same variables were adjusted for confounders at the multivariate level (Table 3), they remained statistically significant (p<0.05), implying that they were factors associated with ocular morbidity among stone quarry workers.

**Table 3:**
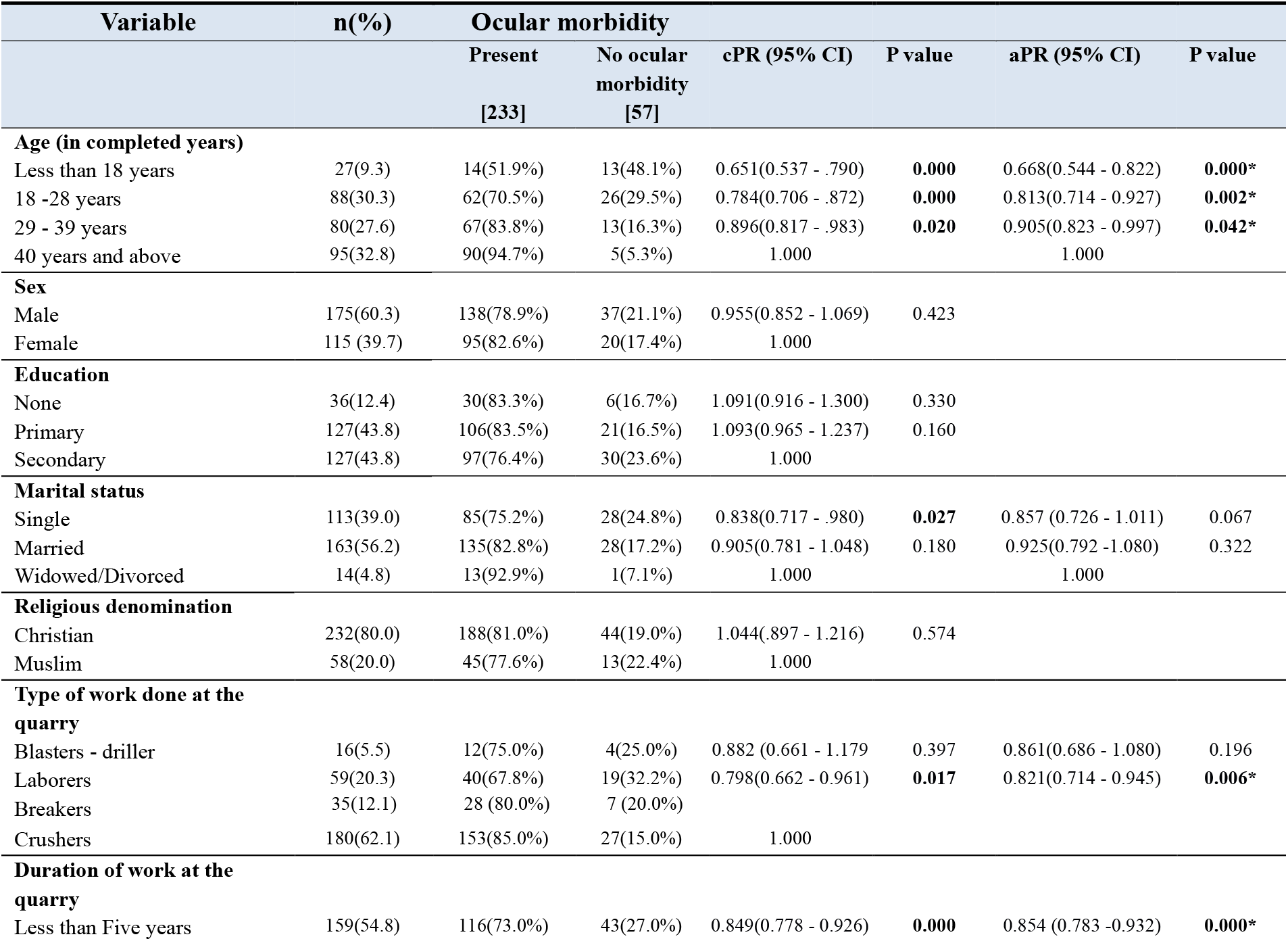
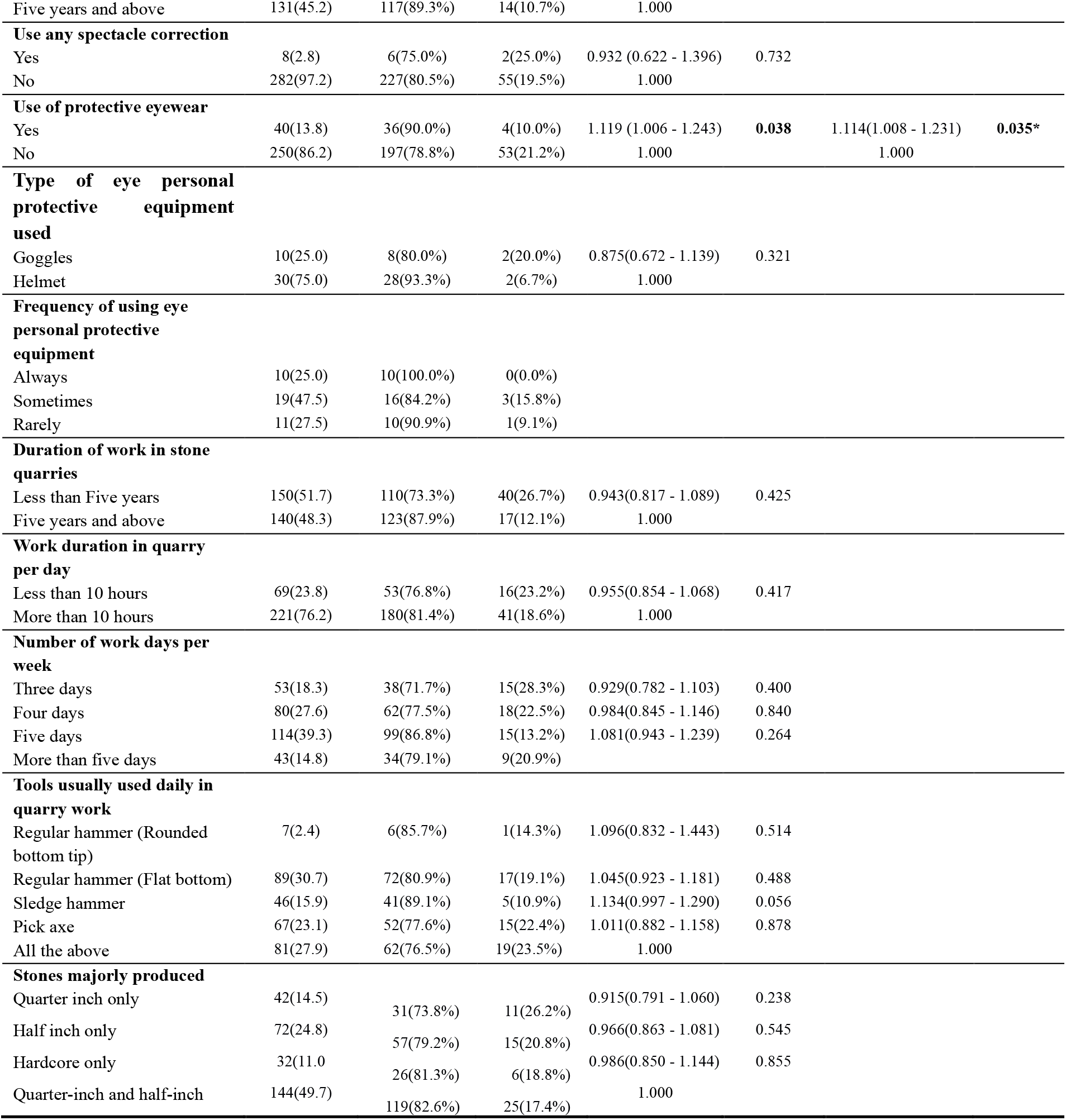
Multivariate analysis of socio-demographic and occupational factors associated with ocular morbidity among stone quarry workers in the districts of Kampala and Mukono, Uganda.

The prevalence of ocular morbidity was 33% lower among stone quarry workers who were younger than 40 years of age (aPR = 0.668 [CI = 0.544 - 0.822], P = 0.000) than among those aged older than 40 years. The prevalence of ocular morbidity was 18% lower among stone quarry workers who were laborers in the quarry (aPR = 0.821 [CI = (0.714 - 0.945], P = 0.006) than among those who were breakers or crushers. The prevalence of ocular morbidity was 15% lower among stone quarry workers who had been working in the quarry for less than five years (aPR = 0.854 [CI = 0.783 -0.932], P = 0.000) than among those who had been working in the quarry for more than five years. The prevalence of ocular morbidity was 11% greater among stone quarry workers who reported wearing protective eyes (aPR = 1.114 [CI = 1.008 - 1.231] p = 0.035*) than among those who did not use protective eyes. Another factor that showed a statistically significant association with ocular morbidity was the non-observation of any workmate suffering vision-threatening eye injuries (Table 4). The findings indicated that the prevalence of ocular morbidity was 19% lower among respondents who agreed that they had seen a workmate suffer severe eye injury due to working in the quarry (aPR = 0.808 [CI = 0.690 - 0.946], p = 0.008).

**Table 4:** Multivariate analysis of work environment-related factors associated with ocular morbidity among stone quarry workers in the districts of Kampala and Mukono, Uganda.

| Variable | n(%) | Ocular morbidity |  | cPR (95% CI) | P value | aPR (95% CI) | P value |
| --- | --- | --- | --- | --- | --- | --- | --- |
|  |  | Present<br>[233] | No ocular morbidity<br>[57] |  |  |  |  |
| Rating of congestion in quarry |  |  |  |  |  |  |  |
| Overcrowded | 63(21.7) | 54(85.7%) | 9(14.3%) | 1.084(0.958 - 1.227) | 0.201 |  |  |
| Somewhat crowded | 142(49.0) | 113(79.6%) | 29(20.4%) | 1.019 (0.913 - 1.139) | 0.732 |  |  |
| Not crowded | 8(29.3) | 66(77.6%) | 19(22.4%) | 1.000 |  |  |  |
| Quarry workers unionized or have an association |  |  |  |  |  |  |  |
| Yes | 118(40.7) | 88(74.6%) | 30(25.4%) | 0.885(0.782 - 1.001) | 0.052 | 0.954(0.873 - 1.041) | 0.289 |
| No | 172(59.3) | 145(84.3%) | 27(15.7%) | 1.000 |  | 1.000 |  |
| Occupation safety rules put in place for workers in quarry, by management |  |  |  |  |  |  |  |
| Yes | 97(33.4) | 72(74.2%) | 25(25.8%) | 0.890 (0.779 - 1.016) | 0.085 | 0.970(0.887 - 1.062) | 0.512 |
| No | 193(66.6) | 161(83.4%) | 32(16.6%) | 1.000 |  | 1.000 |  |
| Frequency of supervision by leaders in this quarry |  |  |  |  |  |  |  |
| Sometimes | 34(11.7) | 26(76.5%) | 8(23.5%) | 0.964(0.784 - 1.185) | 0.725 |  |  |
| Rarely | 130(44.8) | 107(82.3%) | 23(17.7%) | 1.037(0.920 - 1.169) | 0.550 |  |  |
| Never | 126(43.4) | 100(79.4%) | 26(20.6%) | 1.000 |  |  |  |
| Leaders ever educate about occupational safety |  |  |  |  |  |  |  |
| Yes | 125(43.1) | 104(83.2%) | 21(16.8%) | 1.064(0.951 - 1.191) | 0.279 |  |  |
| No | 165(56.9) | 129(78.2%) | 36(21.8%) | 1.000 |  |  |  |
| Fellow workmates in the quarry do not use eye protection |  |  |  |  |  |  |  |
| Disagree | 11(3.8) | 7(63.6%) | 4(36.4%) | 0.781(0.483 - 1.264) | 0.314 |  |  |
| Undecided | 57(19.7) | 48(84.2%) | 9(15.8%) | 1.033(0.836 - 1.278) | 0.761 |  |  |
| Agree | 195(67.2) | 156(80.0%) | 39(20.0%) | 0.982(0.809 - 1.191) | 0.852 |  |  |
| Strongly agree | 27(9.3) | 22(81.5%) | 5(18.5%) | 1.000 |  |  |  |
| Seen a workmate suffer severe eye injury due to working in the quarry |  |  |  |  |  |  |  |
| Disagree | 60(20.7) | 52(86.7%) | 8(13.3%) | 0.940(0.801 - 1.103) | 0.448 | 0.914 (0.773 - 1.081) | 0.293 |
| Undecided | 62(21.4) | 54(87.1%) | 8(12.9%) | 0.944(0.806 - 1.106) | 0.477 | 0.931 (0.792 - 1.095) | 0.389 |
| Agree | 154(53.1) | 114(74.0%) | 40(26.0%) | 0.828(0.712 - 0.964) | **0.015** | 0.808(0.690 - 0.946) | **0.008\*** |
| Strongly agree | 14(4.8) | 13(92.9%) | 1(7.1%) | 1.000 |  | 1.000 |  |
| It is our norm here to work as much as one can for as long as they accumulate a given tonnage of stones |  |  |  |  |  |  |  |
| Disagree | 29(10.0) | 24(82.8%) | 5(17.2%) | 1.007(0.839 -1.209) | 0.940 |  |  |
| Undecided | 97(33.4) | 77(79.4%) | 20(20.6%) | 0.974(0.842 - 1.125) | 0.718 |  |  |
| Agree | 125(43.1) | 100(80.0%) | 25(20.0%) | 0.980(0.852 - 1.126) | 0.773 |  |  |
| Strongly agree | 39(13.4) | 32(82.1%) | 7(17.9%) | 1.000 |  |  |  |
| Workers in this quarry have a high perceived risk of eye injury and disorder |  |  |  |  |  |  |  |
| Disagree | 75(25.9) | 56(74.7%) | 19(25.3%) | 0.948(0.757 - 1.187) | 0.642 |  |  |
| Undecided | 117(40.3) | 98(83.8%) | 19(16.2%) | 1.038(0.839 - 1.285) | 0.730 |  |  |
| Agree | 83(28.6) | 67(80.7%) | 16(19.3%) | 1.007(0.809 - 1.254) | 0.949 |  |  |
| Strongly agree | 15(5.2) | 12(80.0%) | 3(20.0%) | 1.000 |  |  |  |
| Frequency of flooding in specific work areas |  |  |  |  |  |  |  |
| Sometimes | 166(57.2) | 129(77.7%) | 37(22.3%) | 0.933(0.801 - 1.087) | 0.375 |  |  |
| Rarely | 98(33.8) | 82(83.7%) | 16(16.3%) | 0.991(0.847 - 1.159) | 0.906 |  |  |
| Never | 26(9.0) | 22(84.6%) | 4(15.4%) | 1.000 |  |  |  |

## DISCUSSION

From this study, we found that stone quarrying is a male-dominated trade, with most of the workers being married and possessing some form of education. This finding is comparable to that of a study performed in Nigeria (n= 410), in which 82.4% were male [9], but differs from that of another study performed in Nigeria (n=384), in which the percentage of females was 58.9% [3]. This can be explained by the tedious nature of the work involved in stone mining, which includes heavy lifting, the use of explosives, heavy machinery, and the use of rudimentary tools such as sledgehammers.

In this study, the overall prevalence of ocular morbidity among stone quarry workers was found to be very high at 80%, which is comparable to the 78% prevalence found in a study performed in Nigeria [3]. This is attributable to the nature of work, environmental factors in the quarry, and poor/improper use of PPE while working. A study performed in Uganda among small-scale welders revealed that the overall prevalence of ocular disorders among this population was 59.88%, which is significantly lower than that in this study [10]. This difference can be attributed to the level of PPE usage in both groups, where PPE usage among small-scale welders was greater (89%) than that among stone quarry workers in this study (13.8%).

The most prevalent ocular anterior segment disorders in the present study were pinguecula (38.2%), pterygium (34.1%), presbyopia (28.2%), conjunctival hyperpigmentation (20.6%), and dry eye (9.6%). The observed pattern of eye disorders correlates with the observations from several studies [11, 12, 13] where pinguecula was also a leading disorder. The overall high prevalence of pinguecula and pterygium, both of which are degenerative conjunctival disorders, could be attributed to the outdoor work environment and inherent exposure to ultraviolet radiation and dust. Furthermore, the lack of environmental dust control measures and workers’ non-utilization of protective eye devices while at work can exacerbate these conditions [5].

Presbyopia was found in 28.2% of the participants, 32.8% of whom were over 40 years of age, and nearly none of them (97.2%) used spectacle correction. This finding is comparable but significantly lower than that of another study carried out in Nigeria, where presbyopia was observed in 45.5% of the stone quarry workers, with only 30.3% of them wearing near correction [14]. Presbyopia is a disorder associated with aging, and thus, the observed disparities arising from differences in participant and industry characteristics, for example, workers from quarries aged 21-30 years and others from stone processing plants aged 31-40 years, were studied in the Nigerian study [3]. However, in this study, nearly one-third of the population studied was aged 40 years and above, 95 (32.8%).

Conjunctival hyperpigmentation was also a significant finding in this study (20.6%, 60 participants). This finding is characteristic of continuous/long-term exposure to allergens in the environment and is commonly observed in people with allergic conjunctivitis. Other studies [3] categorized this finding as conjunctivitis, which accounted for 13.7% (70 participants) of anterior segment ocular findings in a similar population.

Dry eye was found in 28 participants (9.6%), which is significantly lower than in a study carried out in Egypt among marble quarry workers [5], where the prevalence of dry eye was found to be 60.4%. The factors associated with dry eye in the Egyptian study included age above 40 years, smoking, and a work duration of more than 20 years in the marble processing plant [5], which were probable contributors to the disparity in our study, where the majority of the workers were younger than 40 years of age and had worked for less than 5 years in the quarry.

Cataracts were found in 17 participants, accounting for 4.5% of the participants. This is comparable to a study carried out in Nigeria among stone quarry workers [3] in which the prevalence of cataracts was 6.5%. This could be attributed to the effects of outdoor work-related chronic exposure to ultraviolet radiation and work-related ocular trauma.

Corneal abnormalities, including keratitis, abrasions, corneal scars, corneal foreign bodies, and corneal neovascularization, were found in 13 participants. All these effects were directly attributed to circumstances in the work environment and had a positive correlation with previous ocular injury while at work.

Retinal abnormalities were found in 10 patients (3.4%), which is slightly greater than in a study performed in Nigeria, where retinal pathology contributed only 1.3% [14]. In our study, retinal findings were mainly present in persons aged 40 years and above with other comorbidities, such as HIV, hypertension and diabetes mellitus, while others had no identifiable risk factors.

The prevalence of ocular morbidity was 33% lower among stone quarry workers who were younger than 40 years of age than among those aged 40 years and older. This can be due to the greater risk for injury among the older age group, mainly from inattention and easy fatigability [15]. Another probable explanation is the high prevalence of ocular disorders, such as dry eye disease, presbyopia, conjunctival degeneration, and retinal disorders, among people aged 40 years and older [16].

The prevalence of ocular morbidity was 18% lower among stone quarry workers who were laborers in the quarry than among those who were breakers or crushers. This can be attributed to the nature of the work in these subgroups. Breakers and crushers are at increased risk due to the crushing of stone, which often involves the use of rudimentary tools such as chisels, hammers and pick axes. This puts them in direct contact with stone splinters, dust, flying debris and, occasionally, direct injury from the tools being used. Laborers, on the other hand, are involved in lifting, loading stones and ferrying stones from one point to another [17].

The prevalence of ocular morbidity was 15% lower among stone quarry workers who had been working in the quarry for less than five years than among those who had been working in the quarry for more than five years. This can be explained by the fact that working in open areas for extended periods in the absence of PPE means being exposed to macroclimatic conditions and solar radiation affecting both the ocular surface and anterior segment, thus increasing one’s risk of ocular morbidity [18].

The prevalence of ocular morbidity was 11% greater among stone quarry workers who reported wearing protective eyes than among those who did not use protective eyes. These findings were similar to those of another study carried out among welders in Katwe, Kampala [10], which pointed to a greater likelihood of ocular disorders among welders who sustained injuries while purportedly using protective eyewear. This can be attributed to the improper or occasional use of PPE while working coupled with the failure to take the necessary caution exhibited by those who never use PPE.

The prevalence of ocular morbidity was 19% lower among respondents who agreed that they had seen a workmate suffer severe eye injury due to working in the quarry despite the majority of the participants (154 [53.1%]) agreeing that not seeing any of their workmates suffer a threatening eye injury due to working in the quarry. This finding can be attributed to the behavioral modifications and precautions taken by those who have witnessed a colleague suffer vision-threatening eye injury [19].

### Strengths and limitations of the study

This is the first study on the prevalence, pattern and factors associated with ocular morbidity among stone quarry workers in an East African population. The ocular examinations were cross-checked by a three-person team, reducing intra-observer bias.

However, this study may have information bias, which can include either recall or interviewer bias, which can affect the accuracy of the data obtained, especially those related to accidents sustained at the quarry. Furthermore, there is a lack of pre-employment ocular health records to serve as a baseline for comparison in light of the current findings.

### Conclusion and Recommendations

The prevalence of ocular morbidity among stone quarry workers in the districts of Kampala and Mukono, Uganda, was as high as 80%, with the most common ocular disorders being pinguecula, pterygium, presbyopia, and dry eye. The significantly associated factors included being aged 40 years or older, the type of work engaged in at the quarry (breakers and crushers had increased risk), working at the quarry (study site) for more than 5 years, inconsistent usage of protective eyewear, and no observation of any workmate suffering life-threatening eye injuries.

Routine screening of stone quarry workers for ocular morbidities is recommended. This facilitates early diagnosis and prompt management of vision-threatening ocular morbidities.

Employers need to provide workers with PPE and ensure its consistent and proper usage through routine supervision at the quarry site.

Campaigns by the Ministry of Gender Labor and Social Development (MGLSD) to raise awareness among employers and workers at risk of occupational health hazards within the quarries and how to mitigate them.

Enforcement of the OSH Act by relevant stakeholders to mitigate ocular morbidity among stone quarry workers.

## Data Availability

The data used during this study are available upon request from the corresponding author.

https://makir.mak.ac.ug/search?query=Ahebwa%20Amelia

## ABBREVIATIONS

CDC: Center for Disease Control
IAPB: International Agency for the Prevention of Blindness
ILO: International Labor Organization
LMIC: Low- and Middle-Income Country
MGLSD: Ministry of Gender Labor and Social Development
MOH: Ministry of Health
OSH: Occupational Safety and Health
PPE: Personal Protective Equipment
SOMREC: School of Medicine Research Ethics Committee
WHO: World Health Organization

## DECLARATIONS

### Ethical Approval and Consent to Participate

Ethical approval for the study was obtained from the School of Medicine Research and Ethics Committee (SOMREC) of Makerere University. Additionally, permission to conduct the study was obtained from the Department of Ophthalmology, the Uganda Stone Quarries Association, and the management of the respective stone quarries studied. Before enrollment, written informed consent was obtained from participants, and parental consent/assent was obtained for children aged 17 years and younger. The study was conducted in accordance with the Declaration of Helsinki and the Uganda National Council of Science and Technology guidelines for Human Subject Research.

### Consent for publication

Yes

### Availability of data and materials

The data used during this study are available upon request from the corresponding author.

### Authors’ contributions

AA designed the study, participated in the data collection and statistical analyses, and wrote the first draft of the manuscript. AA supported the data collection and analysis. OJ and AI actively supervised all stages of the study, including manuscript preparation. All the authors have read and approved the final manuscript.

## Acknowledgments

The authors thank the stone quarry workers who participated in the study, including the management of the various stone quarries studied. Heartfelt gratitude is also expressed toward the Department of Ophthalmology lecturers and staff for their commitment.

## Footnotes

1 Required as part of manuscript submission to be included as summary table at end of publication if accepted. Not initially part of the original manuscript write-up.

